# Optimizing Audit and feedbaCk To Implement eVidence-based prAcTices in primary health carE in Nepal, Mozambique, Tanzania and China (ACTIVATE trial): rationale and design of a factorial randomized trial

**DOI:** 10.1101/2025.11.11.25339563

**Authors:** Huanyuan Luo, Hamada Kidanga Mussa, Xiaoqing Zhu, Qing Zhao, Yiyuan Cai, Dadong Wu, Jiangyun Chen, Xiaohui Wang, Changchang Li, Duolao Wang, Mengjun Zhang, Run (Sherry) Wang, Pramesh Koju, Sony Shakya Shrestha, Rajeev Shrestha, Biraj Man Karmacharya, Eusébio Eugênio Chaquisse, Rosa Marlene Cuco, Elsa Luís Kanduma, Isaias Ramiro, Archana Shrestha, Sophia Siu Chee Chan, Dong (Roman) Xu

## Abstract

**Background:** The ability of primary health workers (PHWs) to practice in accordance with evidence-based guidelines and norms is a critical component of improving the quality of primary healthcare. Implementation science seeks to promote the routine use of evidence-based practices in healthcare by identifying barriers and facilitators to their implementation, and by developing strategies such as audit and feedback (AnF) to overcome these barriers. However, the effects of AnF show significant heterogeneity across studies, so this research focuses on the validity of AnF components and the optimal combinations of these components through head-to-head comparisons.

**Methods:** During the preparation phase, we will conduct a preliminary exploration to optimize AnF intervention aimed at improving the quality of primary healthcare. This will involve identifying key candidate components and levels of the AnF intervention, understanding the mechanisms of action, and assessing the resource constraints associated with implementing AnF in primary healthcare (PHC) facilities. This will be achieved through expert consultation and a Best-Worst Scaling (BWS) questionnaire survey. In the optimization phase, we will use the gold standard method of Unannounced Standardized Patients (USP) to assess the quality of primary healthcare for diabetes and hypertension, focusing on the accuracy and standardization of consultation, examination, diagnosis, and treatment procedures in comparison to evidence-based practices, across Nepal, Mozambique, Tanzania, and China. A factorial design randomized controlled trial will be conducted to determine how feedback on care quality can be delivered to PHWs in order to optimize its impact on healthcare quality improvement. To achieve this, the factorial trial will incorporate four key AnF components, each at two levels, resulting in a total of sixteen intervention groups. PHWs will be randomly assigned to these groups.

**Discussion:** This study will provide an empirical foundation for using AnF to improve the quality of primary healthcare in developing countries, while also enhancing and complementing the existing AnF theory. This study focuses on the optimization of the important implementation strategy of AnF in implementation science, contributing a universal research paradigm to the field.

**Trial registration:** ClinicalTrials.gov, NCT06480487. Registered 27 December 2024, https://clinicaltrials.gov/study/NCT06480487.

## Introduction

### Background and rationale

Numerous studies have highlighted the need to improve the quality of primary healthcare (PHC) in low- and middle-income countries (LMICs). Since 2017, our research team has been conducting the *Primary heAlth Care quAlity Cohort In ChinA* (ACACIA) study, using Unannounced Standardized Patients (USPs) to covertly visit more than 2,000 randomly selected PHC facilities across seven provinces of China, in order to gather accurate information on the quality of services. Interim results from the ACACIA study indicate that PHC services in China perform moderately well in terms of efficiency and timeliness of service delivery. However, there are significant concerns regarding the technical quality (the efficacy and safety of diagnosis and treatment) and the quality of patient-centered services. For eleven common diseases, the correct diagnosis rate among primary health workers (PHWs) was only 27.3%, and the standardized rates for consultation, examination, and treatment were only 16.0%, 10.0%, and 23.0%, respectively^1^. These findings suggest that the quality of PHC in China is comparable to that in other LMICs^2–4^.

Implementation science seeks to promote the routine use of evidence-based practices in healthcare by investigating the barriers and facilitators to their implementation, and developing strategies to overcome these barriers. Over time, implementation science has evolved to introduce new concepts aimed at advancing evidence-based practices in PHC, such as radical incrementalism (including Multi-phase Optimization Strategy, or MOST)^6^, Consolidated Framework for Implementation Research (CFIR)^7^, and Reach, Effectiveness, Adoption, Implementation, and Maintenance (RE-AIM)^8^. The MOST is an innovative, systematic approach designed to assist researchers in the development, optimization, and implementation of complex interventions. Its goal is to identify the most effective combinations of intervention components to achieve optimal implementation outcomes, while considering resource constraints. In general, compared to traditional health interventions, there are currently few randomized controlled trials (RCTs) focused on implementation strategies, and even fewer conducted in LMICs.

There are five major types of supply-side implementation strategies aimed at improving healthcare quality: (1) audit and feedback (AnF), (2) training and education, (3) infrastructure improvement (e.g., updating electronic medical record systems), (4) financial incentives, and (5) clinical decision support systems (e.g., implementing physician-specific “reminder” systems)^9, 10^. Training and education for medical staff are among the most commonly used strategies to improve healthcare quality^11, 12^. However, due to the well-known “know-do” gaps in healthcare training and education, the knowledge and skills acquired are not always reflected in actual practice^13, 14^. Infrastructure improvements and financial incentives, while effective in some cases, are not always feasible^15–17^. Clinical decision support systems, including “reminder” mechanisms, have been widely studied for their efficacy but are often of limited value for replication^18^. Therefore, the AnF implementation strategy is the primary focus of this research.

Through AnF, professionals are encouraged to modify their clinical practices to align with evidence-based standards^19^. An audit involves assessing professional performance against explicit criteria or standards, such as evidence-based practices and clinical guidelines. The results of this assessment are then systematically communicated back to the professionals in a structured manner. AnF is a crucial implementation strategy for improving PHC. However, the effects of AnF show significant heterogeneity across studies. This variability may stem from the complexity of AnF components or inconsistencies in its implementation due to vague definitions. Meta-analyses have indicated that the overall effect size of AnF as an intervention was established after the first 30 RCTs^20^. However, comparisons of different AnF component combinations primarily rely on indirect comparisons across studies. These comparisons often yield weak evidence-based conclusions due to variations in study designs, settings, intervention types, target behaviour complexities, and outcome indicators^14^. Thus, future research need to prioritize evaluating the validity of individual AnF components and determining the optimal combination of these components through direct, head-to-head comparisons.

Additionally, there is a lack of theoretical foundations guiding the establishment of components in existing AnF research. According to the literature, few AnF studies explicitly utilize theory to inform their development, implementation, and evaluation^21^. Instead, the majority of studies rely primarily on the researchers’ experience or intuition.

The accuracy of the audit is the foundation of effective feedback. Chart review is the most commonly used audit method. While chart review audits are straightforward to implement, they are often imperfect in PHC settings in LMICs. Information may be omitted, over-recorded, or inaccurately documented, meaning the results of chart reviews do not necessarily reflect the true quality of PHC. In recent years, Unannounced Standardized Patients (USPs) have been increasingly adopted for assessing PHC quality^1^. USPs are highly trained individuals who can realistically and consistently simulate the symptomatic and emotional characteristics of specific conditions. By following standard healthcare service procedures without informing the healthcare providers of their role, USPs can capture a realistic and comprehensive picture of the quality of care delivered. However, to our knowledge, no studies to date have utilized USPs in the context of AnF.

Up to 140 RCTs on AnF have been conducted^22^, with the majority taking place in developed countries such as the United States, Canada, the United Kingdom, Sweden, Australia, and New Zealand^23–25^. To the best of our knowledge, only a few studies have been conducted in China and LMICs^26–29^ and none in Mozambique, Tanzania, and Nepal. It is necessary to develop and validate the optimal AnF interventions for LMICs in PHC settings.

### Objectives

This study aims to identify the most effective combination of AnF components, taking into account resource limitations, to enhance the implementation of clinical guidelines and improve the quality of PHC in Mozambique, Zanzibar (Tanzania), Nepal, and China. Specifically, the study seeks to determine how the results of quality of care obtained from the audit process can be effectively fed back to healthcare providers in the context of PHC in LMICs, thereby optimizing improvements in care quality. Guided by the MOST strategy, we aim to accomplish the following three goals: (1) To assess the quality of the consultation process, examination, diagnosis, and treatment provided by PHC facilities in the four LMICs using USPs tool; (2) To explore the components and resource constraints (or optimization criteria, such as cost and time) for the AnF intervention in the four countries; and (3) To identify the optimal AnF intervention packages tailored to local contexts by evaluating the main and interaction effects of components, considering optimization criteria specific to PHC settings.

## Methods

This protocol is reported according to the Standard Protocol Items: Recommendations for Interventional Trials (SPIRIT) statement (Additional file 1).

### Preparation for AnF intervention

During the preparation phase, we will conduct a preliminary exploration of how AnF for quality of care can be optimized for healthcare workers, with the goal of enhancing healthcare quality. This will involve identifying key candidate components and levels of the AnF intervention, understanding the mechanisms of action, and assessing the resource constraints associated with implementing AnF in PHC facilities. The findings from this preparation phase will serve as the foundation for the optimization phase, where further investigation will focus on refining the effectiveness of AnF for quality improvement. The details of the preparation for the AnF intervention are as follows.

### AnF component ranking by expert meetings and Best-Worst Scaling (BWS)

Researchers and experts will convene to identify the most important AnF intervention components (such as the seven key components) and levels derived from the literature (Additional file 2). The research team will present the study using PowerPoint slides outlining the potential AnF components, providing experts with an overview of their tasks. These tasks will include discussing the importance, feasibility, and mechanisms of the identified components and levels, as well as ranking their importance and feasibility. The meeting will be recorded using Tencent Conference Meeting software.

Based on the most important AnF components identified during expert meetings, a Best-Worst Scaling (BWS) survey will be used to invite a broader range of stakeholders to rank the relative importance of the AnF candidate components and identify the key components. The BWS survey is a screening experiment grounded in random utility theory. By prompting participants to identify the best and worst options among a series of components, the questionnaire triggers a trade-off mechanism, allowing for the quantification of the relative importance of each component and helping to identify the most crucial one. Its efficiency in reducing the number of paired comparisons, coupled with its strong ability to maintain consistency in judgments, has made it a widely used and effective multi-criteria decision-making technique.

The Balanced Incomplete Block Design (BIBD) is an experimental design used in BWS to enhance results by organizing items into blocks and balancing the frequency of item presentations across participants. This approach allows for efficient comparison of a set of items, ensuring that each item is presented an equal number of times across blocks. The BIBD minimizes bias and improves the statistical precision of the ratings. It is particularly useful when dealing with a larger number of ranked items. For example, BIBD could be implemented in the standard manner by dividing seven AnF intervention components into seven blocks, each containing three components (Table 1). Each participant will receive a questionnaire with all seven blocks. Participants will be asked to assess which component from each block they consider most effective and which they consider least effective.

**Table 1:** BWS questionnaire example.

| Components | Most effective | Least effective |
| --- | --- | --- |
| Both verbal and written feedback | ☐ | ☐ |
| Feedback reports incorporate comparisons with peer data | ☐ | ☐ |
| Feedback reports provide practitioners with suggested actions to overcome barriers (Action implementation toolkit: e.g. customized training manuals) | ☐ | ☐ |

Participants in the BWS survey will include the study population of the trial, as well as administrators. While there are some guidelines, we find no universal standard exists for determining an adequate sample size for BWS surveys^33^. The sample sizes in BWS studies have ranged from 15 to 803 participants, with a median of 175^34^. Based on this, we have decided to invite 180 participants to complete the formal survey. To administer the survey, we will recruit four investigators responsible for distributing and collecting the questionnaires, either online or onsite. The Principal Investigator (PI) and a research assistant will train the investigators on the study objectives and how to administer the questionnaire. The investigators will coordinate the survey logistics, including the time and location for the onsite survey.

Both the counting approach and logit modelling approach will be used for statistical analysis. The counting approach will be employed to calculate the best-minus-worst (B-W) scores, representing the difference between the number of times a given component is selected as “most effective” and “least effective”. A positive B-W score indicates that the component is chosen as “most effective” more frequently than as “least effective”. Logit models will be used to estimate the coefficients for each component, identifying the most and least effective components in promoting greater completion of guideline-recommended items and ensuring accurate diagnosis and treatment by practitioners. The key AnF components will be selected for the optimization of the AnF intervention.

### Study design - factorial RCT

In the optimization phase, a 2×2×2×2 factorial design RCT will be conducted, incorporating four two-level components decided from BWS survey and consensus meeting, resulting in a total of sixteen AnF intervention groups. The PHC workers, policymakers, and health system administrators will be invited to discuss the feasibility and operational details of implementing the sixteen AnF interventions for PHC facilities. Participants will be randomly assigned to one of these sixteen AnF intervention groups. The audited quality of care results will be fed back to the healthcare workers, and outcome indicators reflecting the effectiveness of AnF will be measured. We will estimate the main and interaction effects of AnF components on improving PHC quality via statistical analysis. The optimal AnF combination will be determined by evaluating both the effects and resource constraints in local implementation settings. We assume that the four AnF components in Table 2, each with two levels, will be selected from the BWS survey. The intervention setup is presented in Table 2.

**Table 2:** Intervention groups.

| Group | Component 1 (Delivery Method of Feedback) |  | Component 2 (Source of Feedback) |  | Component 3 (Feedback with Peer Comparison) |  | Component 4 (Feedback with Goals) |  |
| --- | --- | --- | --- | --- | --- | --- | --- | --- |
|  | Level 1 | Level 2 | Level 1 | Level 2 | Level 1 | Level 2 | Level 1 | Level 2 |
|  | Provided face to face | Provided by electronic mail | Provided by researchers | Provided by authoritative bodies | Provided with peer comparison | Provided without peer comparison | Provided with goals | Provided without goals |
| 1 | ✓ |  | ✓ |  | ✓ |  | ✓ |  |
| 2 |  | ✓ | ✓ |  | ✓ |  | ✓ |  |
| 3 | ✓ |  |  | ✓ | ✓ |  | ✓ |  |
| 4 | ✓ |  | ✓ |  |  | ✓ | ✓ |  |
| 5 | ✓ |  | ✓ |  | ✓ |  |  | ✓ |
| 6 |  | ✓ |  | ✓ | ✓ |  | ✓ |  |
| 7 |  |  |  |  |  |  |  |  |
| 8 | ✓ |  |  | ✓ |  | ✓ | ✓ |  |
| 9 | ✓ |  | ✓ |  |  | ✓ |  | ✓ |
| 10 |  | ✓ | ✓ |  |  | ✓ | ✓ |  |
| 11 |  | ✓ | ✓ |  | ✓ |  |  | ✓ |
| 12 | ✓ |  |  | ✓ |  | ✓ | ✓ |  |
| 13 | ✓ |  |  | ✓ | ✓ |  |  | ✓ |
| 14 | ✓ |  | ✓ |  |  | ✓ |  | ✓ |
| 15 |  | ✓ |  | ✓ |  | ✓ | ✓ |  |
| 16 | ✓ |  |  | ✓ |  | ✓ |  | ✓ |

### Study sites

The study sites will include Maputo City and Maputo Province in Mozambique; the North, South, and Urban West regions in Zanzibar, Tanzania; the Kathmandu, Bhaktapur, and Lalitpur districts in Nepal; and Henan Province, Guangdong Province, and other regions in China.

## Study population

### Inclusion and exclusion criteria

In Mozambique, the inclusion criteria are nurses, general medicine technicians, and physicians in PHC facilities; and the exclusion criteria are healthcare providers from other departments, such as tuberculosis, malaria, human immunodeficiency virus/acquired immune deficiency syndrome, or emergency services, as well as students currently on internship. In Zanzibar, Tanzania, the inclusion criteria are clinical officers, nurses, and other healthcare providers involved in consultation, examination, diagnosis, and treatment in PHC facilities. In Nepal, the inclusion criteria are medical officers, health assistants, and senior auxiliary health workers in PHC facilities; and the exclusion criteria are interns or students present during the visit. In China, the inclusion criteria of PHC facilities are primary and secondary hospitals, community health centers (stations) and clinics, as well as township health centers and village health clinics; and the inclusion criteria of research participants are practicing (assistant) physicians and rural physicians working in healthcare facilities that meet the above criteria, with a scope of practice only including general practice and internal medicine.

### Sample size

The sample of the 2×2×2×2 factorial design study is calculated based on the primary outcome: the completion rate and adherence to key consultation items in guidelines by workers at PHC facilities (measured as a continuous variable). As stated in the introduction, the ACACIA project, conducted by our research group in China, reported a mean completion rate of key consultation items of 16.0% (standard deviation = 12.7%) from interim data, which is comparable to that observed in other LMICs. Preliminary expert consensus suggests that an increase of approximately 5 percentage points in the completion rate (effect size: unstandardized mean difference) is considered clinically significant. The parameter for the standard deviation of the response variable after treatment, within each treatment condition (i.e., adjusting for treatment but not for post-test), is not available in the literature. Therefore, we use the available data from the ACACIA project. We consider a significance level of 0.05 (two-sides) and a power of 0.9. Using the “FactorialPowerPlan()” command in the MOST package of R software (version 4.4.2), the total sample size required is 274. To ensure equal numbers in each intervention group, we plan to recruit 18 participants per group. Considering potential subject attrition and other factors that may result in a dropout rate of approximately 5%, the adjusted sample size for each group is 19 participants, resulting in a total sample size of 304.

### Variables, data collection instrument and outcome

Variables collected at baseline include facility characteristics (i.e., country, diseases managed by USP [diabetes or hypertension]), and PHC workers characteristics (i.e., age, gender, education level, years of work experience).

The study will focus on Type II diabetes and hypertension, as these are prevalent conditions in primary care. Outcome indicators will be systematically collected following the RE-AIM framework and Proctor’s Implementation Outcomes Framework^8^. Among these indicators, the quality of care indicators will be assessed using the USP methodology and subsequently provided as feedback to PHC workers. The quality of care indicators will align with the quality framework developed by the Institute of Medicine (IOM)^30^, which consists of six domains: effectiveness, efficiency, equity, patient-centeredness, safety, and timeliness.

- **Effectiveness** (avoiding underuse and misuse) and **safety** (avoiding harm) represent the traditional technical goals of quality care. These encompass aspects such as consultation, examination, diagnosis, and treatment, and will include both the primary and some secondary outcomes.
- **Patient-centeredness** (respecting and accommodating individual preferences) will be evaluated using the *Patient Perception of Patient-Centeredness Rating Scale* (PPPC)^31^. The PPPC employs a 4-point Likert scale to measure attitudes across three dimensions: exploring the illness and the experience of being ill, understanding the whole person, and finding common ground.
- **Timeliness** will be assessed through measures such as facility opening hours, waiting times, and consultation durations.

Efficiency (avoiding waste) can be evaluated by analyzing the costs incurred for care provided to the USPs during their interactions with PHC workers. Equity (ensuring no disparities in quality due to individual characteristics) cannot be fully evaluated through a single USP visit and will require a separate study. Due to funding and time limitations, both the efficiency and equity will not be assessed in this study.

The primary outcome is the proportion of completed guideline-recommended quality checklist items for consultation of hypertension and Type II diabetes cases of the PHC providers among all of the items. This outcome will be expressed as a continuous score ranging from 0% to 100%.

Secondary outcomes include:

- Quality of care indicators: The proportion of completed guideline-recommended quality checklist items for physical and laboratory exams of hypertension and Type II diabetes cases of the PHC providers among all of the items. This is a continuous score ranging from 0 to 100%.
- Quality of care indicators: Correctness of diagnosis of hypertension and type II diabetes cases by PHC providers. This will be categorized into 2 categories: correct and incorrect diagnosis.
- Quality of care indicators: Correctness of treatment of hypertension and type II diabetes cases by PHC providers. This will be categorized into 2 categories: correct and incorrect treatment.
- Quality of care indicators: Timeliness of hypertension and type II diabetes services in PHC settings.
- Quality of care indicators: Patient-centered quality of healthcare in PHC settings.
- Implementation outcome: Adoption of AnF intervention by study participants. It is a dichotomous variable, consisting of two categories: adopted, not adopted. It will be self-reported by participants using team-developed questionnaire.
- Implementation outcome: Costs to researchers of developing and implementing AnF intervention. It is a continuous variable and will be assessed using project final account of expenditure.
- Implementation outcome: Participants score of acceptability of AnF intervention. It will be self-reported by participants using Generic Theoretical Framework of Acceptability (TFA)-based questionnaire.

### Procedure of randomization and data collection

Step 1: Informed consent/basic information/audit. Study participants will be invited to complete the informed consent form and basic information questionnaire on line or offline. USPs seek medical care without disclosing their identity as USPs, following standard procedures to gather information on quality of care indicators. All data will be entered into REDCap^32^, a free and robust cloud-based tool for data collection, storage, and management. The auditing content (i.e., quality of care indicators) will be collected using the *Quality of Care Assessment Form* for later feedback. This data will be documented either by the USPs themselves or by investigators after conducting interviews with the USPs completing their visit to the participants.

Step 2: Randomization and intervention. Following the completion of the basic information questionnaire by all study participants, randomization will be performed. The randomization lists will be generated using block randomization with a block size of sixteen. Eligible participants will be randomly assigned to one of the sixteen AnF intervention groups, with stratification by country. The randomization process will be carried out by two statisticians who are independent of the trial. They will perform the assignment sequentially to ensure accuracy and minimize bias. Given the nature of the intervention content (e.g., the intervening physicians would receive feedback reports from the program coordinator), it is not possible to blind the study coordinator or participating physicians to the intervention assignments. However, outcome assessors and data analysts are not aware of these assignments.

Step 3: Collection of outcome indicators after intervention completion. For the outcomes related to quality of care indicators, we will use USP tools for data collection, following a method similar to the baseline audit. For implementation outcomes, including the adoption of the AnF intervention by study participants and participants’ acceptability scores of the AnF intervention, we will conduct a survey to gather the data.

### Data analysis and management

Statistical significance will be assessed using a two-sided alpha of 0.05. All statistical analyses will be performed using SAS software version 9.4. The participant baseline characteristics, and primary and secondary outcomes, will be shown as means (standard deviation) or number (proportions) as appropriate. We will conduct the intention-to-treat (ITT) analysis, i.e. the analysis will be conducted based on the originally assigned groups. We will use generalized linear models (GLMs) to evaluate the effectiveness of the intervention. In this study, the primary outcome is the proportion of completed guideline-recommended quality checklist items for consultation of hypertension and Type II diabetes cases of the PHC providers among all of the items, which is a continuous score. For the primary outcome, we will employ a GLM with a normal distribution and a identity link function, from which mean differences and its 95% confidence interval (95% CI) will be derived.

For sensitivity analysis, the missing values of the primary outcome will be imputed using multiple imputation method to verify the robustness of the primary analysis. We will also conduct subgroup analyses, using GLMs to examine the intervention’s effectiveness on primary outcome across various characteristics, including country, diseases managed by USP [diabetes or hypertension], and PHC workers characteristics (i.e., age, gender, education level, years of work experience). The economic evaluation of the sixteen interventions will be conducted using the incremental analysis method of cost-effectiveness analysis.

For continuous secondary outcomes, the same statistical analysis methods for analysis of the primary outcome will be applied. For dichotomous secondary outcomes, we will use GLMs with a binomial distribution and an identity link function, from which risk differences and its 95% CI will be derived.

## Discussion

This study will provide an empirical foundation for using AnF to improve the quality of primary healthcare in developing countries, while also enhancing and complementing the existing AnF theory. This study focuses on the optimization of the important implementation strategy of AnF in implementation science, contributing a universal research paradigm to the field. Unlike previous studies that explore entirely new interventions, or those that seek various solutions to barriers in implementation research, this study applies theories and frameworks of implementation science to optimize the AnF implementation strategy. The expected outcome is that this research paradigm will be applicable to the optimization of implementation strategies with similar research problems in the future, enhancing the intervention effects of these strategies, and thus contributing to the development of research paradigms in implementation science. This, in turn, represents both the challenge and difficulty of this study. Specifically:

First, our study contributes to the existing literature by examining [research topic] across four countries, all classified as LMICs, with some categorized as low-income. While most previous research in this field has been conducted in high-income settings, studies from LMICs remain limited. By addressing this gap, our findings will provide valuable insights into the implementation challenges and strategies within resource-constrained environments. Moreover, the cross-country analysis enhances the generalizability of our results, providing a comparative perspective that highlights both common and context-specific factors influencing AnF effectiveness. These insights are particularly relevant for informing policy and practice in similar settings globally.

Second, regarding the concept of intervention development, the study follows the guidance of the radical incremental approach. The radical incremental approach is a commonly used concept in intervention development in implementation science, emphasizing the gradual and stepwise improvement of intervention measures during the development process to achieve a “radical” enhancement of intervention effectiveness. For complex intervention packages, which consist of multiple components aimed at facilitating implementation (e.g., AnF strategy), combining the most effective components can yield the most efficient overall intervention package. This approach reduces the risk of developing ineffective interventions compared to the development of entirely new interventions.

Third, in terms of the development process, the study adopts the principle of “co-production”. By closely collaborating with stakeholders (using the Delphi expert consultation method and BWS survey), the study jointly develops and tests the AnF intervention. This approach promotes communication and improves the development efficiency, effectiveness, feasibility, and sustainability of the intervention.

Fourth, in terms of research focus, the study centers on the components and combinations of intervention components. Previous studies on the AnF effectiveness have shown considerable variation, potentially due to the differences in the components and combinations of interventions under the general term of AnF. There has been a lack of investigation into whether the AnF components and their combinations are effective or exhibit synergistic effects. Therefore, this study focuses on identifying which components of the AnF implementation strategy are effective in improving the quality of primary healthcare of diabetes and hypertension (primarily concerning the accuracy and standardization of procedures for consultation, examination, diagnosis, and treatment), as well as the effects of these components when combined.

Finally, in terms of research design, a Multi-phase Optimization Strategy is employed. This research framework is used for the development, optimization, and evaluation of complex intervention packages. In contrast to previous studies that used parallel designs, comparing multiple groups with different feedback methods or comparing them with a control group, the advantage of the multi-phase optimization strategy is as follows: 1) It uses an efficient factorial design (with a smaller sample size to validate more components) to explore whether each component of the intervention is effective and to examine potential synergistic effects between components, such as whether the combined effects strengthen or weaken the overall impact; 2) It emphasizes “optimization under constraints”, taking into account local resource limitations to achieve the most effective AnF implementation strategy.

## Data Availability

All data produced in the present work are contained in the manuscript.

## Abbreviations

PHWs: Primary health workers
AnF: Audit and feedback
PHC: Primary healthcare
BWS: Best-Worst Scaling
USP: Unannounced Standardized Patients
LMICs: Low- and middle-income countries
ACACIA: *Primary heAlth Care quAlity Cohort In ChinA*
MOST: Multi-phase Optimization Strategy
CFIR: Consolidated Framework for Implementation Research
RE-AI: Reach, Effectiveness, Adoption, Implementation, and Maintenance
RCTs: Randomized controlled trials
BIBD: Balanced Incomplete Block Design ITT Intention-to-treat
GLMs: Generalized linear models
CI: Confidence interval

## Authors’ contributions

HL, HKM, XZ, and DX designed the study. HL and DX secured funding. HL drafted the manuscript. QZ, YC, SSS, RS, BMK, EEC, RMC, ELK, and IR contributed to the design of the intervention and the development of the implementation approach. DW, JC, XW, CL, DL, MZ, RW, PK, AS, and SSCC contributed to the development of the study methodologies and protocols. The authors reviewed and approved the final manuscript.

## Funding

This study is funded by the National Natural Science Foundation of China (72404118), China Postdoctoral Science Foundation (2023M731536), Guangdong Medical Science and Technology Research Foundation (A2023133), and Swiss Agency for Development and Cooperation (81067392).

## Availability of data and materials

Not applicable.

## Declarations

### Ethics approval and consent to participate

This study obtained ethics approval under a protocol approved by the relevant authorities in China, Nepal, Mozambique and Zanzibar, Tanzania. All participants will provide written informed consent.

### Competing interests

The authors declare no competing interests.

## Notes

### Competing Interest Statement

The authors have declared no competing interest.

### Clinical Trial

NCT06480487

### Funding Statement

This study is funded by the National Natural Science Foundation of China (72404118), the China Postdoctoral Science Foundation (2023M731536), the Guangdong Medical Science and Technology Research Foundation (A2023133), and the Swiss Agency for Development and Cooperation (81067392).

