## Additional file 2 for "Optimizing Audit and feedbaCk To Implement eVidence-based prAcTices in primary health carE in Nepal, Mozambique, Tanzania and China (ACTIVATE trial): rationale and design of a factorial randomized trial"

**Additional file 2: Audit and feedback (AnF) components**

| S.N | Components | Levels | Description |
| --- | --- | --- | --- |
| 1 | Feedback with peer comparison | 1. Yes | In addition to the feedback, the feedback report will include a comparison with peers/other health institutions. There will be a comparison between healthcare facilities (PHCs) regarding clinical performance/quality of care/patient-centered health service for the management of hypertension and diabetes. (Your current adherence rate of management of diabetes as per the Desk guide for non-communicable diseases and risk factors is 60%, whereas the adherence rate of management of diabetes of Mwera health facility is 75%, and Marumbi health facility is 80%.) |
|  |  | 2. No |  |
| 2 | Feedback with goals/targets | 1. Yes | In addition to feedback, the feedback report will include the goals/targets to be achieved regarding clinical performance/quality of care/patient-centered health service for the management of hypertension and diabetes within a certain time period. (e.g.Your current adherence rate of the standard management of diabetes as per Desk guide for non-communicable diseases and risk factors is 60%. You have to increase it to 80% in next 3 months) |
|  |  | 2. No |  |
| 3 | Feedback with an improvement plan | 1. Yes | In addition to feedback, the feedback report will include an improvement plan for the healthcare workers. There will be suggestions for improvements and action plans for diabetes and hypertension management. |
|  |  | 2. No |  |
| 4 | Feedback with clinical guidelines | 1. Yes | In addition to feedback, we will provide a standard treatment protocol (e.g. Desk guide for non-communicable diseases and risk factors**)** for the management of hypertension and diabetes for an educational purpose. |
|  |  | 2. No |  |
| 5 | Feedback with hazard information | 1. Yes | In addition to feedback, the feedback report will include a hazard information alert to healthcare workers regarding hypertension and diabetes. E.g. "Unregulated hypertension and diabetes care may lead to 1) increased cardiovascular risk; 2) kidney impairment; 3) vision problems; 4) neurological problems; 5) difficulty with blood glucose control; 6) Medication side effects; 7) Increased health care costs." |
|  |  | 2. No |  |
| 6 | Feedback with patient voice | 1. Yes | In addition to feedback, the feedback report will include the voice of patients, i.e. the quality of diabetes and hypertension services experienced, expected, and desired by the diabetic and hypertensive patients / USPs. |
|  |  | 2. No |  |
| 7 | Delivery method of feedback report | 1. Face-to-face (Handing feedback report physically) | The feedback provider will visit the health institution and deliver the feedback face-to-face to feedback recipients. |
|  |  | 2. Mails (Electronic) | The feedback provider will provide feedback through electronic mail to the feedback recipients. |
|  |  | 3. Phone calls (Electronic) | Feedback providers will provide feedback through phone calls to the feedback recipients. |
|  |  | 4. Text messages through social media (Electronic) | The feedback provider will provide feedback through text message to the feedback recipients. |
| 8 | Format of the feedback report | 1. Textual report (Feedback report is in texts only) | Feedback reports will be only in textual format. There will be no visual displays such as graphs, or charts. For e.g. "Your completion rate of consultation, completion rate of examination, diagnostic accuracy, and treatment accuracy for this hypertension diagnosis were 60%, 40%, 40%, and 40% respectively". |
|  |  | 2. Feedback report designed with texts and visual displays (graphs, figures and charts) | The feedback report will be in the textual and visual display format. Overall Consultation Completion Rate Charts/Graphs: Use bar charts and graphs to demonstrate how well healthcare professionals are performing clinically in implementing hypertension and diabetes guidelines. |
| 9 | Feedback framing | 1. Positive Framing | The sentence structure of feedback is framed in such a way that it reflects the positive aspects of the performance. (E.g. Your accuracy of diagnosis is 60%) |
|  |  | 2. Negative Framing | The sentence structure of feedback is framed in such a way that it reflects the negative aspects of the performance. (e.g. Your diagnosis is 40% inaccurate) |
|  |  | 3. Mixed Framing | The sentence structure of feedback is framed in such a way that it reflects both positive aspects and negative aspects of the performance. |
| 10 | Source of feedback | 1. Authoritative bodies | For example, monitoring & evaluation supervision bodies like the Health section of municipalities, districts, and provinces directly notify the health care provider about their clinical performance for the month. |
|  |  | 2. Researcher | Those researchers who were part of the audit will directly notify primary care physicians of his clinical presentation this month. |
| 11 | Feedback recipients | 1. Clinician who manages the diabetes/hypertension patients (Whom we audit the quality of care) | Health care worker who is responsible for the diabetes and hypertension case management. |
|  |  | 2. Clinician and management team members of that health facility | Feedback recipients will be both healthcare workers and the management team members of that health facility |
| 12 | Response to feedback | 1. Provision to get responses to feedback from recipients | For example, a primary care worker who receives feedback, responds to the researchers with his/her comments about the feedback intervention. |
|  |  | 2. Provision not to get responses to feedback from recipients | Primary care worker who receives feedback will not respond to the researchers with his/her comments about the feedback intervention. |
| 13 | Frequency of feedback intervention | 1. One time | Feedback will be provided only once. |
|  |  | 2. Multiple times | Feedback will be provided more than once. |
| 14 | Reminder messages | 1. Sending reminder messages every week for 3 months | We will send reminder messages to the recipients of the feedback every week for 3 months |
|  |  | 2. Sending reminder messages monthly for 3 months | We will send reminder messages to the recipients of the feedback every month for 3 months |
|  |  | 3. Not sending a reminder message | We will not send reminder messages to the recipients of feedback |
